# Tool for Implementing TArget Trial emulatioN (TITAN): an open-access design assistant

**DOI:** 10.1101/2025.11.07.25339253

**Authors:** Malamati Voulgaridou, Isabelle Boutron, Raphaël Porcher, Viet-Thi Tran, Elodie Perrodeau, Anna Pellat, Philippe Ravaud, Sally Yaacoub

**Author notes:** **Corresponding author:** Sally Yaacoub, BS Pharm, MPH, PhD, Université Paris Cité and Université Sorbonne Paris Nord, Inserm, INRAe, Center for Research in Epidemiology and Statistics (CRESS), Cochrane France, Hôpital Hôtel Dieu, 1 place du Parvis Notre-Dame, 75004 Paris, France.

## Abstract

**Background:** Target trial emulation offers a structured framework to reduce bias in observational studies by emulating the design of a hypothetical target randomised controlled trial. Despite a growing interest for this framework, the implementation of target trial emulation is often inconsistent, with design flaws frequently introducing avoidable biases.

**Objective:** To develop a design assistant tool to provide structured guidance to researchers for planning and conducting target trial emulations.

**Methods:** A multi-disciplinary steering committee defined the scope, content and structure of Tool for Implementing TArget Trial emulatioNs (TITAN). First, we reviewed the literature to identify concepts according to the steps of planning and designing a target trial emulation. Then, we developed the tool through an iterative process with two rounds of pilot testing and subsequent refinement by the steering committee.

**Results:** TITAN provides guidance to emulate a pragmatic two-arm trial assessing a pharmacological intervention. The tool is organized in two main sections. First, the users define the research question and the target trial. Then, they emulate the target trial using the available observational data. The tool provides warnings, in addition to suggestions, to minimize avoidable biases and major methodological errors; particularly focusing on the alignment of study time points (eligibility, treatment assignment, and start of follow-up) and related biases. The tool’s output is a synthesis that includes a summary of the target trial, design of its emulation, and possible biases and solutions.

**Conclusions:** TITAN could help researchers ensure that observational studies follow the key principles of target trial emulation.

## 1. Introduction

Observational studies can play a crucial role in informing decisions about pharmacological treatment effectiveness in real-world settings. Compared to randomised controlled trials (RCTs), they can generate evidence more quickly and at lower cost than RCTs, and are the only source of evidence when RCTs are infeasible (1). Observational studies assessing the effectiveness of interventions are nevertheless prone to several biases (2).

Many of these biases can be avoidable when they arise from suboptimal design choices (3). In particular, they are often caused by the misalignment of three key study time points: eligibility (when patients fulfil the eligibility criteria), treatment assignment (when patients are assigned to one of the treatment strategies), and start of follow-up (when data start to be considered to evaluate the outcome). Failure to align these time points can impose a risk of biases in effect estimates, such as selection bias and immortal time bias (3). Similarly, confounding bias, resulting from failure to identify or adequately account for confounding factors, should be mitigated. Therefore, a way to ensure that observational studies preserve some methodological features of randomised trials is to design such studies so that they explicitly emulate a hypothetical RCT (i.e., target trial) that would answer the research question. Target trial emulation aims to prevent common design and analytical errors in observational studies that could otherwise lead to misleading causal inferences (4).

Interest in target trial emulation has increased significantly in recent years, as reflected by the growing number of published studies using this approach (5). Although the theoretical framework for target trial emulation is well-established, its reporting has been inconsistent, prompting the development of TARGET reporting guideline (4, 6). Similar inconsistencies exist for the framework’s practical implementation. A study has shown that approximately 50% of published target trial emulations have time point misalignment, stemming from design flaws rather than limitations of the data itself (7). Such methodological issues could be addressed early in the study planning phase and common design errors and biases may be avoided.

Hence, we developed Tool for Implementing TArget Trial emulatioNs (TITAN), an open-access web-based design assistant that can be used in the planning of observational studies aiming to emulate a target trial assessing a pharmacological intervention, in order to minimize avoidable biases and major methodological errors. The tool can be accessed here: https://clinicalepidemio.fr/target_trial/

### Objective

Our objective was to develop an online open-access design assistant tool to provide structured guidance for researchers for planning and conducting target trial emulations, in order to minimize avoidable biases and major methodological errors.

## 2. Methods

We adopted a multi-step approach for the development of TITAN as detailed below.

### Steering committee

We set up a steering committee composed of six experts in the field of research methodology and target trial emulations, including a biostatistician (RP) and epidemiologists (MV, SY, VTT, PR, IB). The steering committee defined the scope, content and structure of the tool, as well as refined the tool after pilot-testing.

### Scope of the tool

TITAN was intended to be used for designing observational studies emulating a target trial, that would be: 1) pragmatic – i.e. having less stringent eligibility criteria, comparing an intervention and a comparator arm that are more flexibly defined as it would be in clinical practice – as recommended in the target trial emulation framework (8); 2) assessing a pharmacological intervention, and more specifically the initiation and/or adherence to the intervention; and 3) comparing two arms of intervention. Studies comparing treatment strategies (e.g. durations of treatment, sequences of treatments), or comparing three or more interventions, were not considered in the current version of the tool.

The target audience of TITAN are researchers conducting target trial emulations. It was designed to support users who may have limited expertise in causal inference and target trial emulation by providing step-by-step guidance in defining the research question and designing a target trial emulation. Nonetheless, it does not replace the expertise of methodologists or statisticians, which is necessary for the conduct of robust observational studies.

### Content of the tool

Through a literature review, we identified key papers related to the target trial framework (8), the importance of the synchronisation of the time points in observational studies (9, 10), as well as recognized tools to design, assess or report observational studies, such as STaRT-RWE (11), ROBINS-I (12) and TARGET (6). We extracted the key concepts identified, related to the biases by design and main challenges that arise during the process of target trial emulation. Then, we developed a visual mapping of all the identified concepts, where we organized them according to the steps of planning and designing a target trial emulation (miro.com/app/board/uXjVJagi5z4=/). Building on the visual mapping, the steering committee defined the tool’s content and structure, through iterative meetings and discussions. The final content and structure of the tool were approved by the steering committee.

### Pilot-testing of the tool

The tool was pilot-tested in two rounds. The primary version of the tool was tested by three users, including a clinician (AP), a biostatistician (EP) and an epidemiologist (GKRB). They went through the content and structure of the tool, in order to assess the overall understanding of the presented concepts. The feedback and suggestions of the users informed the subsequent revisions of the tool.

After iterative revisions, the final version of the tool was pilot-tested by four users, including two members of the steering committee (RP, VTT), a clinician (AP), and an epidemiologist (AD). They pilot-tested the tool using a published RCT, depending on their specialty, as the target trial, and attempting to emulate the trial using a data source of their choice. In order to ensure the applicability and usability of the tool, users provided feedback on its content, overall structure, clarity, and ease of completion, as well as identified possible technical errors of the online version.

## 3. Results

The tool is organized in two main sections: I. Definition of the target trial and II. Design of emulation using observational data. The output is a synthesis presenting a summary of the target trial and the emulation, and a supplementary section providing additional information on the target trial emulation framework and the statistical methods proposed throughout the tool.

First, the users define the research question and the target trial (RCT), then they operationalize the definitions using observational data in order to emulate the target trial (i.e., map the study elements to observational data). We focus on the alignment of the study time points and biases by design, and provide warnings and suggestions, in order to avoid biases by design and major methodological errors. The structure and flow of the tool are presented in Figure 1.

**Figure 1.**
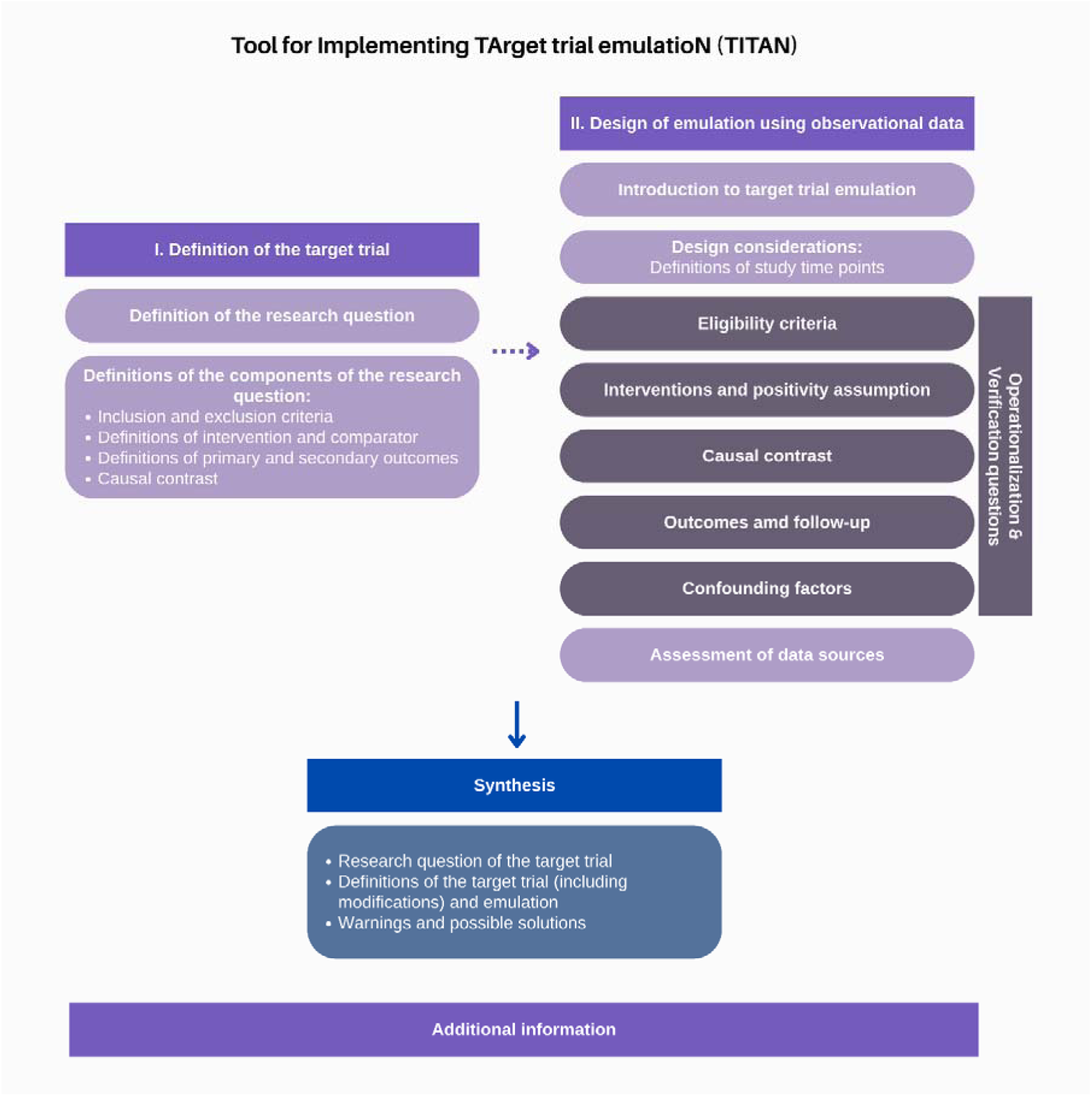
TITAN overall structure

### I. Definition of the target trial

#### Definition of the research question

Users are first asked to clearly define the research question of the target trial by using the PICO (Population, Intervention, Comparator, Outcomes) framework. A well-defined research question is essential for any study (13) and particularly critical in observational studies, where the availability of data may influence or bias the formulation of the research question.

#### Definitions of the components of the research question

Users are asked to specify each eligibility criterion (inclusion and exclusion), define the intervention and comparator, and define the primary and secondary outcomes to be analysed (e.g., measurement variable and time point), considering the pragmatic nature of the trial. In addition, users are asked to define the causal contrast of the target trial, that is, the comparative effect of the treatment strategies evaluated. Although multiple options are described in the literature (14), we focus on the following two:

- Effect of assignment to intervention (Intention to treat effect): measuring the effect based on the initial assignment to a treatment, whether or not the treatment was fully followed during the study (12, 15). It is also referred to as ‘treatment policy strategy’ (14).
- Effect of adherence to intervention: measuring the effect based on starting and adhering to the assigned treatment throughout the study (12, 15). It is also referred to as ‘while on treatment strategies’ (14).

These components of the study are the cornerstone of the causal estimand of RCTs (14). An estimand is a precise description of the treatment effect reflecting the clinical question underlying the trial objective. Once defined, a trial can be appropriately designed to facilitate a reliable estimation of the targeted treatment effect (14).

### II. Design of emulation using observational data

#### Introduction

We introduce the target trial emulation framework highlighting the importance of aligning the time points of the study (16), as presented in the tool (see Figure 2):

- Time point of eligibility: when participants will be considered eligible for inclusion
- Time point of treatment assignment: when participants will be classified to one of the treatment groups; and
- Time point of start of follow-up: when the assessment of outcomes begins.

**Figure 2.**
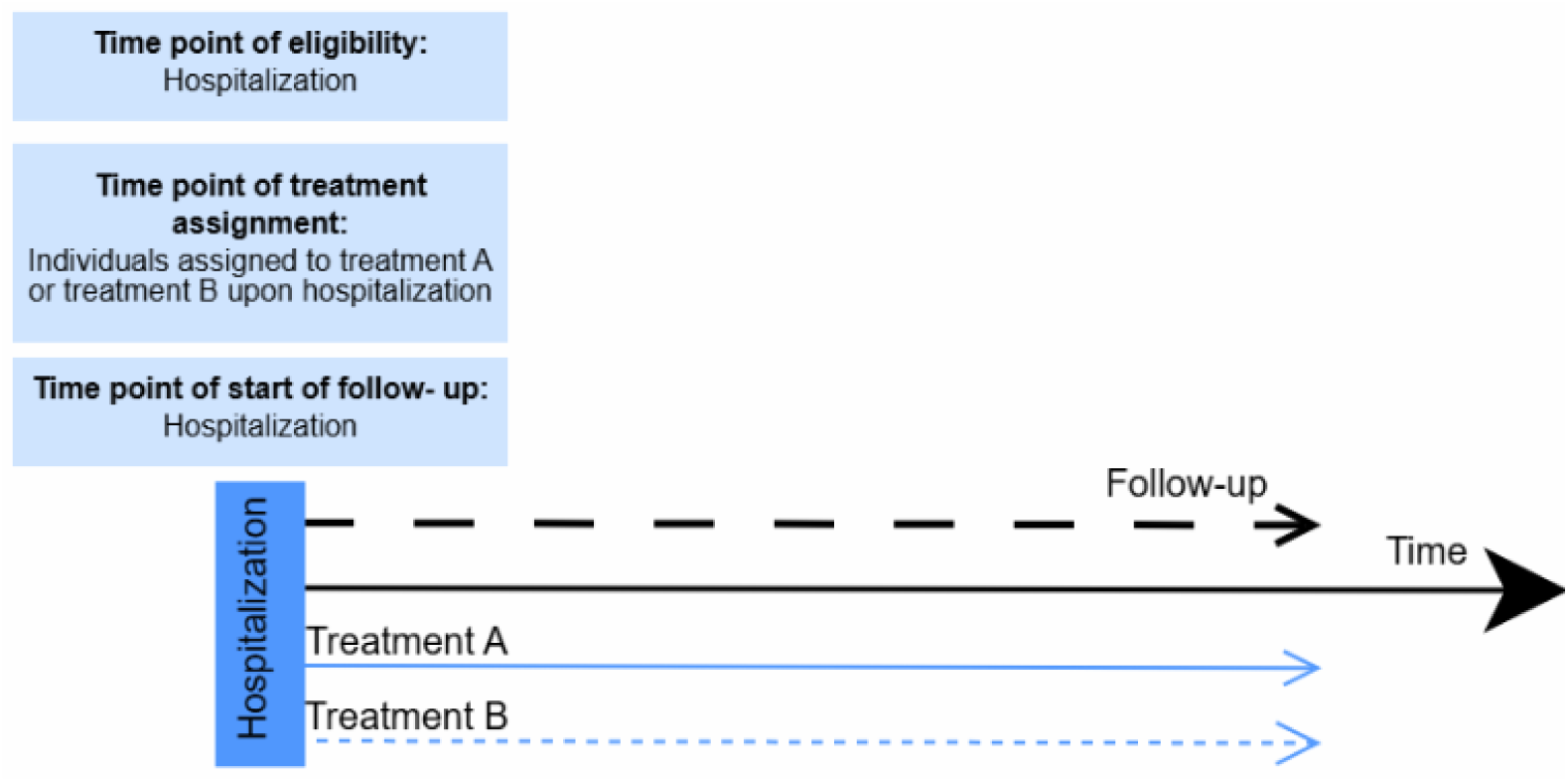
Example of a diagram illustrating the alignment of the three time points

Additionally, a triggering event is defined as a clinical situation when a treatment decision is made and helps in defining the time points. For example, hospitalization for myocardial infarction can prompt considering prescription of treatment A or not. In the absence of a triggering event, participants can be eligible at multiple times. For example, a target trial comparing initiation versus no-initiation of hormonal therapy in postmenopausal women on the risk of breast cancer. Initiation of treatment could occur at any time after the diagnosis of menopause until she starts treatment or has the outcome of interest (breast cancer) (8).

The time points of eligibility, treatment assignment and start of follow-up are usually aligned in RCTs (16) : participants meet the criteria for inclusion at the same time point of assignment to the treatment group, while the assessment of their outcomes simultaneously begins. Therefore, the lack of synchronization, or misalignment, of the time points can lead to different biases. For example, when the assessment of the outcomes (such as the collection of their clinical outcomes) begins at a later time than their inclusion to the study and the treatment initiation, then the time point of start of follow-up is misaligned with those of eligibility and treatment assignment. Another example is when there is a grace period (i.e., a delay between eligibility and treatment initiation during which participants are not yet assigned to a treatment group). If present, a grace period should be defined and addressed using statistical methods such as the clone-censor-weight approach. This involves creating ‘clones’ of individuals, assigning them to the different strategies of the emulated trial, and censoring them based on the observed actual treatment received (8, 9), in order to minimize immortal time bias and misclassification of treatment. For example, in a target trial emulation assessing the impact of beta-blockers on mortality and cardiovascular outcomes in patients with obstructive sleep apnoea, a 12-month grace period existed after the time point of eligibility, during which the treatment initiation was possible, and subsequently addressed this using the clone-censor-weight approach (17).

Other possible situations of time point misalignment exist as well (9).

#### Design considerations

Users are asked whether there is a triggering event for the treatment decision. For example, myocardial infarction is a triggering event to decide whether or not to treat patients with beta-blockers. If there is no triggering event, then users participants may be eligible at multiple times (until they start treatment, or until they develop the outcome assessed), therefore, users should decide to either conduct one trial by choosing one of the multiple eligibility times, or conduct a series of trials.

Then, users are asked to define each of the time points of their study, as the lack of time point alignment can lead to avoidable biases, as presented in Box 1 (18).

##### Box 1 Biases by design due to time point misalignment

- Prevalent user bias: a type of selection bias that occurs when individuals already taking the treatment (i.e., prevalent users) are included, rather than those who are now starting it (new users or incident users). This bias arises because prevalent users have already survived the early phase of treatment, during which adverse effects or other outcomes may have occurred, thereby underestimating risks related to the intervention (12). The time point of treatment assignment precedes that of start of follow-up.
- Selection bias due to post-treatment eligibility: which occurs when participants are selected based on criteria applied after treatment initiation, leading to systematic exclusion of individuals who fail to meet post-treatment conditions, such as receiving a minimum number of prescriptions or surviving a specific period after treatment initiation (18). The time point of treatment assignment precedes that of start of follow-up.
- Immortal time bias: arise when a period of the follow-up is either excluded (i.e. not accounted for in the analysis) or misclassified (i.e. participants are not classified into the correct treatment arm). The immortal time refers to the interval during which the outcome could not have occurred, since participants must survive to receive the treatment assignment, and thus are considered as “immortal” during this interval (10, 19). The time point of start of follow-up precedes that of treatment assignment.
- Misclassification bias: (or exposure misclassification) occurs when during the aforementioned interval of immortal time, it is unclear to which treatment group the participants should be assigned to (18).

#### Data sources

Users are asked to specify their data sources (such as electronic health records or claims data), as it will dictate the emulation of the study.

#### Operationalization

In the tool’s first section, users define a target trial that is pragmatic and addresses a relevant clinical question, rather than one that is driven by the available data. In this second section, users are asked to verify if their data allows for the identification of the study elements, i.e., to operationalize the study elements of their target trial (14). However, we recognize that emulating a target trial can be challenging or even infeasible depending on the availability of the data. In such cases, slight changes in the target trial might render the emulation feasible. Therefore, we allow users to generate a modified target trial i.e., one that can reasonably be emulated with the available observational data (6). For transparency, we capture the modified target trial alongside the operationalization (i.e., identification or mapping) of the study elements using the observational data.

Then, users are asked a series of verification questions regarding their study design, coupled with relevant explanations and examples (Box 2). According to the answers provided, warnings and respective solutions are proposed to the users, in order to minimize commonly occurring biases. The methods suggested are non-exhaustive, and the consultation of a statistician during the process of the target trial emulation is strongly encouraged.

##### Eligibility criteria

Users are presented with a summary of the inclusion and exclusion criteria of the target trial and are asked to operationalize them using the selected data source(s); by emulating the original criteria directly or by modifying them before operationalization, as explained above (6, 14). For example, in a target trial assessing the comparative effectiveness of two SGLT2i medications on cardiovascular events, the eligibility criteria of the target trial included patients with type 2 diabetes on any antihyperglycemic treatment, including insulin. However, the operationalization of the eligibility criteria was done through the identification of dispensations of antihyperglycemic medications and the data source did not provide differentiation between type 1 and 2 diabetes for the indication regarding insulin treatment. Therefore, the eligibility criteria were modified and any dispensation of insulin treatment was excluded (20).

Afterwards, users are posed with verification questions to detect avoidable biases (see Box 2), regarding the inclusion of participants based on future events, a minimum number of prescriptions or a specific duration of treatment required for eligibility, or questions regarding prevalent users - all of which might lead to avoidable biases, if not addressed.

###### Box 2 Examples of verification questions to detect avoidable biases and respective proposed solutions

ELIGIBILITY CRITERIA

- Do the eligibility criteria include any “future” events, i.e. events that occur after the start of treatment?
  - Warning: risk of selection bias due to post-treatment eligibility
  - Proposed solution: reconsideration of the eligibility criteria
- Did the participants have to receive the treatment for a specific duration or receive a minimum number of prescriptions in order to be considered eligible or to be classified into the intervention group?
  - If required to establish eligibility
    - Warning: risk of immortal time bias and of selection bias due to post-treatment eligibility
    - Proposed solutions: reconsideration of the eligibility criteria, clone-censor-weight approach
  - If required to be classified into the intervention group
    - Warning: risk of immortal time bias and of misclassification of treatment
    - Proposed solutions: alignment of eligibility and treatment assignment time points, random assignment of participants to treatment groups during the analysis, clone-censor-weight approach
- Are “prevalent users” included, i.e. participants that were receiving the treatment before their follow-up started?
  - Warning: risk of prevalent user bias
  - Proposed solution: reconsideration of the eligibility criteria and exclusion of prevalent users

INTERVENTIONS

- Did you decide to have a delay between the time point of eligibility of participants and the start of treatment, where they are not assigned to either intervention group?
  - Warning: risk of immortal time bias and of misclassification of treatment
  - Proposed solutions: alignment of eligibility and treatment assignment time points, random assignment of participants to treatment groups, clonecensor-weight approach

POSITIVITY ASSUMPTION

- Is the positivity assumption satisfied, i.e., can participants with similar characteristics receive the experimental intervention and comparator during the same time period?
  - Warning: a causal effect cannot be estimated and the emulation of the target trial will not be possible
  - Proposed solutions: design a new study

##### Interventions & Positivity assumption

Similarly, users are presented with the definitions of the intervention and comparator of the target trial and are asked to operationalize them, either by applying the original definitions directly or after modifications. Then, users are asked to verify that the positivity assumption (i.e., the condition that individuals in the study population have a non-zero probability of receiving each treatment option, given their observed characteristics, is satisfied, as it is required for valid causal inference) (21). If the assumption is not satisfied, the warning states that the causal effect cannot be estimated and does not permit the users to advance to further parts of the tool, but rather suggests starting the design of a new study, in order to discourage the design of methodologically flawed target trial emulations (see Box 3). For example, if a target trial assessed amoxicillin versus clarithromycin for the treatment of tonsillitis, and the contraindications of included patients to either treatment were not considered, we would likely be uncertain whether all patients had a non-zero probability of receiving either treatment option. Thus, the positivity assumption would not be verified.

Subsequently, users are asked to specify whether the treatment is time-varying, i.e., referring to patients receiving or not the treatment at various time points over the course of their follow-up, a common phenomenon in real-world practice which requires advanced statistical methods to account for it (22–25). An additional verification question is posed on whether a delay exists between the eligibility and the treatment assignment of participants, during which they are not assigned to either intervention group (see Box 3).

##### Causal contrast, Outcomes and follow-up and Confounding factors

In these sections, users are asked to operationalize the causal contrast of the target trial emulation (6), outcomes and confounding factors considering the data source(s). If this operationalization is not possible, users can specify the modifications for the causal contrast and outcomes and are presented with a warning on the risk of unmeasured confounding in the study coupled with proposed methods for adjustment (e.g., multivariable regression analysis, propensity-based methods, g-computation), when relevant.

#### Completeness and quality of the data source

Finally, users are prompted to explore the data sources used for the emulation of the target trial, in order to identify and address issues that may arise. These include missing data, which might lead to bias due to loss of follow-up, and classification errors, which might lead to classification bias.

### III. Synthesis

After the tool completion, users are provided with a synthesis of their input. This includes a summary of the defined PICO of the target trial, a table illustrating the design elements and the definitions of the target trial, modified target trial and the emulation. In addition, a summary of possible biases detected in the study design as well as proposed solutions is provided. In Figure 3, we present a synthesis generated by TITAN for a target trial emulation of a study assessing beta-blockers after myocardial infarction with preserved left ventricular systolic ejection fraction, inspired by a published study by Matthews et al (26). The original study has been modified for demonstration purposes. The entire synthesis is available Appendix 1.

**Figure 3.**
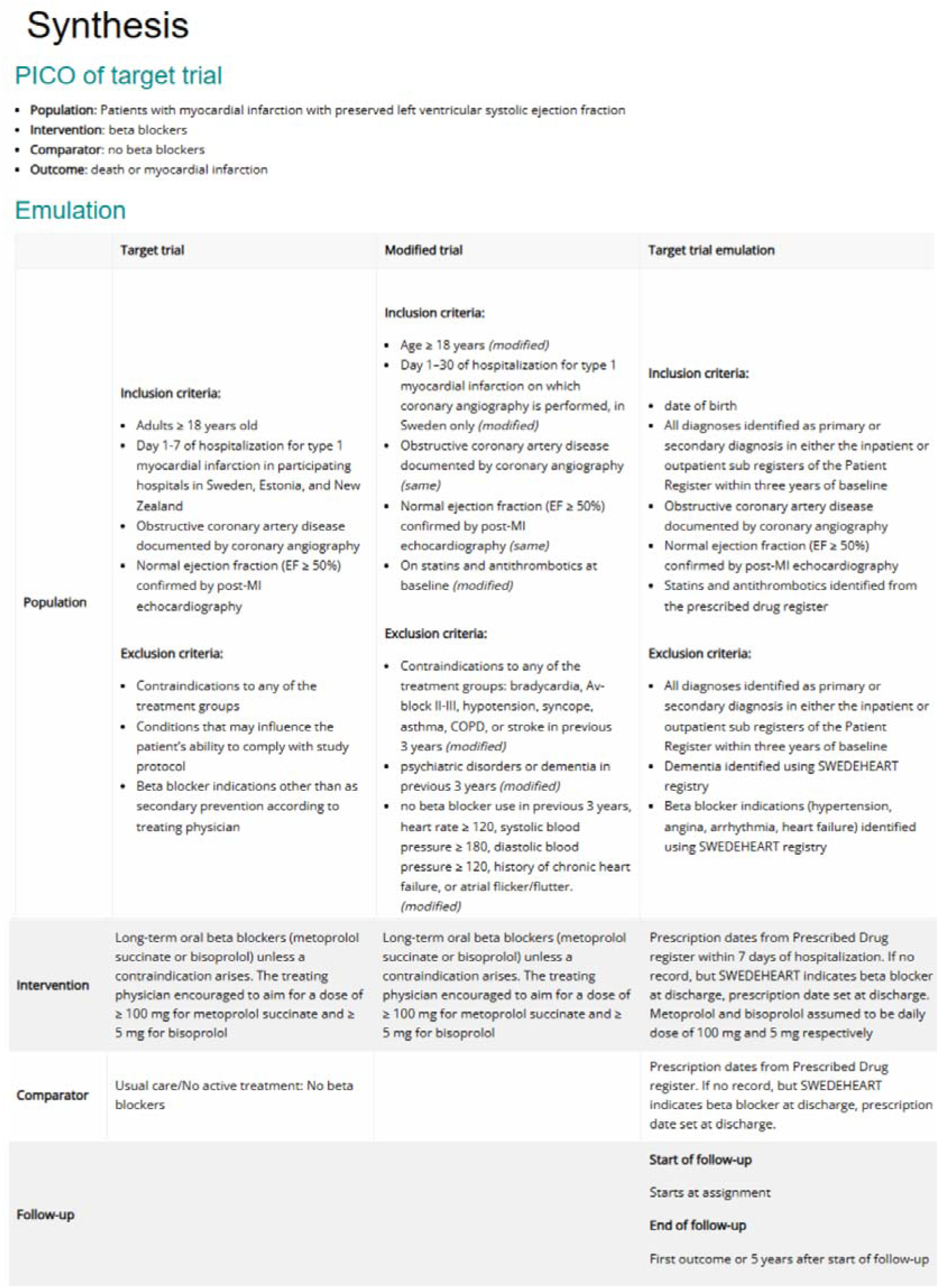
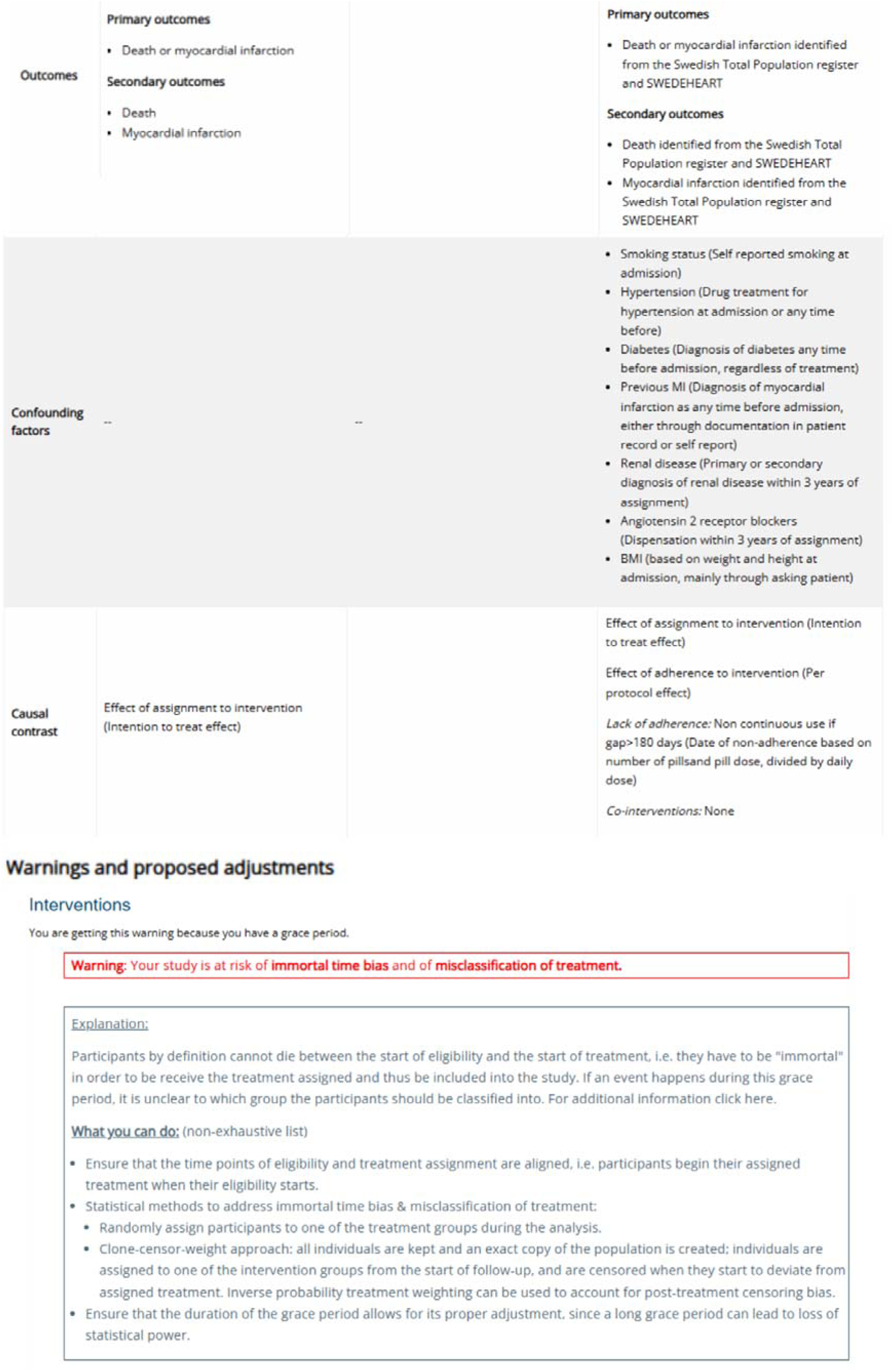
Example of a synthesis generated by TITAN (inspired by Matthews et al.)

## 4. Discussion

The target trial emulation framework can improve the quality of observational analyses by avoiding common methodological pitfalls in the design of observational studies (9). While the framework holds great promise for estimating the causal effect of interventions using observational data, its complexity may limit its adoption by researchers with limited experience in causal inference. TITAN aims to simplify the process of target trial emulations by offering a user-friendly and step-by-step approach. By helping users avoid common errors and biases by design, the tool aspires to enhance the methodological rigor of observational studies emulating a target trial, and therefore their contribution in decision-making in healthcare.

Several other tools for observational studies have been developed, such as the STaRT-RWE, a template for planning and reporting of real-world evidence studies (11), the SPIFD, a structured process assisting in feasibility assessments and identifying fit-for-purpose data for real-world studies (27), and TARGET, a guideline to aid the reporting of these studies (6). While such tools provide valuable frameworks for ameliorating the reporting and data relevancy in observational studies, our tool is designed to actively support users specifically during the planning of the target trial emulation. What differentiates the TITAN is its focus on guiding researchers through the practical steps of target trial emulation, helping to minimize common errors and biases during the study design.

### Strengths and Limitations

TITAN has several strengths. It proposes a structured and systematic approach, providing verification questions, warning messages and guidance along the main steps of planning and designing a target trial emulation. The tool’s revision by experts in the field of research methodology ensures the validity of its content and its alignment with the main methodological issues of observational studies, while the piloting by clinicians guarantees that it remains user-friendly and easy to use. Furthermore, the tool’s online availability makes it easily accessible by users, while its synthesis section provides users with a summary of the design of their emulated trial which can aid them alongside the planning of their study.

Nevertheless, TITAN also has some limitations. The current version of the tool cannot apply to studies aiming to emulate more complex trials such as strategy trials, trials evaluating individualized treatment strategies, or those comparing multiple arms. Similarly, it does not apply to studies assessing non-pharmacological interventions (e.g., surgery, physical rehabilitation) due to the complexity of the intervention and influence and expertise of health care providers. Moreover, while the tool provides essential guidance, it does not substitute the expertise in designing a target trial emulation, where the consultation of a statistician with dedicated expertise remains necessary throughout the design of the study. However, the tool may also help methodologists and statisticians who lack expertise and experience on target trial emulation.

### Future directions

Future directions for TITAN include expanding its applicability beyond its initial scope: specifically, by creating additional versions of the tool, we aim to broaden its applicability beyond pharmacological interventions and refine its guidance to support more complex study designs, accommodating thus a wider range of clinical questions. Additionally, user feedback following the tool’s dissemination will be instrumental in refining its content and enhancing its online version. Potential improvements include clarifying certain methodological aspects based on common user difficulties, providing more concrete examples for each step of the tool, and incorporating additional references to relevant methodological literature to further assist researchers in designing high-quality observational studies.

### Implications

As the use of real-world data continues to expand in healthcare research, tools like TITAN could play a crucial role in improving causal inference and reducing bias in observational studies. By providing a step-by-step guidance for emulating randomised trials, the tool could empower researchers to leverage real-world evidence more effectively, potentially influencing clinical decision-making and thus patient outcomes. Importantly, TITAN can also serve as a decision-support tool, as investigators may deem the study as infeasible or scientifically unjustified, if the risk of bias is too high or the necessary data is not available. TITAN also promotes methodological transparency by requiring users to document the initial study plan as well as any subsequent modifications.

Additionally, TITAN could serve as a valuable educational resource for training researchers and students in the principles of target trial emulation, aiming to strengthen their methodological skills regarding the design of observational studies. Moreover, its systematic approach may support the adoption of standardized methodologies in observational studies and thus encourage more widespread acceptance of observational findings in decision-making.

As the healthcare landscape increasingly relies on real-world evidence, tools like TITAN can facilitate the design of methodologically sound studies, ensuring that observational data contribute meaningfully to evidence-based medicine.

In conclusion, TITAN aims to provide structured guidance for researchers planning to emulate a trial using observational data. Although it does not replace the expertise of methodologists or statisticians, it helps ensure that observational studies follow the key principles of target trial emulation, thus supporting the development of more reliable evidence for drawing valid causal conclusions. By addressing common biases by design and methodological challenges inherent in observational studies, TITAN has the potential to enhance the credibility and validity of such studies and therefore their role in decision-making in healthcare. As the use of real-world data continues to expand, ensuring rigorous methodology in observational research will be essential for the contribution of such studies in comparative effectiveness research.

## Data Availability

No data was used for the research described in the article.

## ACKNOWLEDGEMENTS

We thank Professor Gro Karine Rosvold Berntsen and Professor Antoine Duclos for their contribution in piloting the tool. We thank Ms Elise Diard for developing the online tool.

## DATA STATEMENT

No data was used for the research described in the article.

## AUTHOR CONTRIBUTIONS

MV: Conceptualization, Methodology, Investigation, Writing – original draft

IB: Conceptualization, Methodology, Supervision, Validation, Writing – review and editing

RP: Methodology, Writing – review and editing

VTT: Methodology, Writing – review and editing

EP: Methodology, Writing – review and editing

AP: Methodology, Writing – review and editing

PR: Conceptualization, Validation, Writing – review and editing

SY: Conceptualization, Methodology, Investigation, Supervision, Validation, Writing – original draft

All authors reviewed and approved the final manuscript.

## LIST OF ABBREVIATIONS

PICO: Population, Intervention, Comparator, Outcomes
RCTs: randomised controlled trials
TITAN: Tool for Implementing TArget Trial emulatioN

## SUPPLEMENTARY MATERIAL

**Appendix 1.** Example of a synthesis output

