## Appendix 1 for "Tool for Implementing TArget Trial emulatioN (TITAN): an open-access design assistant"

#### SUPPLEMENTARY MATERIAL

**Appendix 1.** Example of a synthesis output

*Note: Modifications have been applied to the original study of Matthews et al. for demonstration purposes.*

#### Synthesis

Title: Beta blockers vs no beta blockers in patients with myocardial infarction

##### PICO of target trial

**Population:** Patients with myocardial infarction with preserved left ventricular systolic ejection fraction
**Intervention:** beta blockers
**Comparator:** no beta blockers
**Outcome:** death or myocardial infarction

Emulation

| | **Target trial** | **Modified trial** | **Target trial emulation** |
|---|---|---|---|
| **Population** | **Inclusion criteria:**<br>• Adults ≥ 18 years old<br>• Day 1-7 of hospitalization for type 1 myocardial infarction in participating hospitals in Sweden, Estonia, and New Zealand<br>• Obstructive coronary artery disease documented by coronary angiography | **Inclusion criteria:**<br>• Age ≥ 18 years *(same)*<br>• Day 1–30 of hospitalization for type 1 myocardial infarction on which coronary angiography is performed, in Sweden only *(modified)*<br>• Obstructive coronary artery disease documented by coronary angiography *(same)* | **Inclusion criteria:**<br>• Date of birth<br>• All diagnoses identified as primary or secondary diagnosis in either the inpatient or outpatient sub registers of the Patient Register within three years of baseline<br>• Statins and antithrombotics identified from the prescribed drug register<br>**Exclusion criteria:** |

| | | | |
|---|---|---|---|
| | • Normal ejection fraction (EF ≥ 50%) confirmed by post-MI echocardiography<br>**Exclusion criteria:**<br>• Contraindications to any of the treatment groups<br>• Conditions that may influence the patient's ability to comply with study protocol<br>• Beta blocker indications other than as secondary prevention according to treating physician | • Normal ejection fraction (EF ≥ 50%) confirmed by post-MI echocardiography *(same)*<br>• On statins and antithrombotics at baseline *(new)*<br>**Exclusion criteria:**<br>• Contraindications to any of the treatment groups: bradycardia, Av-block II-III, hypotension, syncope, asthma, COPD, or stroke in previous 3 years *(modified)*<br>• psychiatric disorders or dementia in previous 3 years *(modified)*<br>• no beta blocker use in previous 3 years, heart rate ≥ 120, systolic blood pressure ≥ 180, diastolic blood pressure ≥ 120, history of chronic heart failure, or atrial flicker/flutter. *(modified)* | • All diagnoses identified as primary or secondary diagnosis in either the inpatient or outpatient sub registers of the Patient Register within three years of baseline<br>• Dementia identified using SWEDEHEART registry<br>• Beta blocker indications (hypertension, angina, arrhythmia, heart failure) identified using SWEDEHEART registry |
| **Intervention** | Long-term oral beta blockers (metoprolol succinate or bisoprolol) unless a contraindication arises. The treating physician encouraged to aim for a dose of ≥ 100 mg for metoprolol succinate and ≥ 5 mg for bisoprolol | | Prescription dates from Prescribed Drug register. If no record, but SWEDEHEART indicates beta blocker at discharge, prescription date set at discharge. Metoprolol and bisoprolol assumed to be daily dose of 100 mg and 5 mg respectively |
| **Comparator** | Usual care/No active treatment: No beta blockers | | Prescription dates from Prescribed Drug register. If no record, but SWEDEHEART indicates beta blocker at discharge, prescription date set at discharge. |

| | | | |
|---|---|---|---|
| **Follow-up** | | | **Start of follow-up:** At assignment<br>**End of follow-up:** At first outcome or 5 years after start of follow-up |
| **Outcomes** | **Primary outcomes**<br>• Death or myocardial infarction<br>**Secondary outcomes**<br>• Death<br>• Myocardial infarction | | **Primary outcomes**<br>• Death or myocardial infarction identified from the Swedish Total Population register and SWEDEHEART<br>**Secondary outcomes**<br>• Death from the Swedish Total Population register and SWEDEHEART<br>• Myocardial infarction identified from the Swedish Total Population register and SWEDEHEART |

| | | | |
|---|---|---|---|
| **Confounding factors** | -- | -- | • Smoking status (Self reported smoking at admission)<br>• Hypertension (Drug treatment for hypertension at admission or any time before)<br>• Diabetes (Diagnosis of diabetes any time before admission, regardless of treatment)<br>• Previous MI (Diagnosis of MI as any time before admission, either through documentation in patient record or self report)<br>• Renal disease (Primary or secondary diagnosis of renal disease within 3 years of assignment)<br>• Angiotensin 2 receptor blockers (Dispensation within 3 years of assignment)<br>• BMI (based on weight and height at admission, mainly through asking patient) |
| **Causal contrast** | Effect of assignment to intervention (Intention to treat effect) | | Effect of assignment to intervention (Intention to treat effect)<br>Effect of adherence to intervention (Per protocol effect)<br>*Lack of adherence:*<br>Non continuous use if gap>180 days (Date of non-adherence based on number of pills and dose, divided by daily dose)<br>*Co-interventions:*<br>None |

### Warnings and proposed adjustments

#### Interventions

##### Warning:

You are getting this warning because you have a grace period. Your study is at risk of immortal time bias and of misclassification of treatment.

**Explanation:** Participants by definition cannot die between the start of eligibility and the start of treatment, i.e. they have to be "immortal" in order to be receive the treatment assigned and thus be included into the study. If an event happens during this grace period, it is unclear to which group the participants should be classified into.

##### What you can do:

- Ensure that the time points of eligibility and treatment assignment are aligned, i.e. participants begin their assigned treatment when their eligibility starts.
- Statistical methods to address immortal time bias et misclassification of treatment:
    - Randomly assign participants to one of the treatment groups during the analysis.
    - Clone-censor-weight approach: all individuals are kept and an exact copy of the population is created; individuals are assigned to one of the intervention groups from the start of follow-up, and are censored when they start to deviate from assigned treatment. Inverse probability treatment weighting can be used to account for post-treatment censoring bias.
- Ensure that the duration of the grace period allows for its proper adjustment, since a long grace period can lead to loss of statistical power.
